# Use of respirator vs. surgical masks in healthcare personnel and its impact on SARS-CoV-2 acquisition – a prospective multicentre cohort study

**DOI:** 10.1101/2021.05.30.21258080

**Authors:** Sabine Haller, Sabine Güsewell, Thomas Egger, Giulia Scanferla, Reto Thoma, Onicio B. Leal-Neto, Domenica Flury, Angela Brucher, Eva Lemmenmeier, J. Carsten Möller, Philip Rieder, Markus Ruetti, Reto Stocker, Danielle Vuichard-Gysin, Benedikt Wiggli, Ulrike Besold, Stefan P. Kuster, Allison McGeer, Lorenz Risch, Matthias Schlegel, Andrée Friedl, Pietro Vernazza, Christian R. Kahlert, Philipp Kohler

**Author notes:** Contributed equally (Christian R. Kahlert, Philipp Kohler). Corresponding author: Philipp Kohler, MD MSc, Division of Infectious Diseases and Hospital Epidemiology, Cantonal Hospital St. Gallen, Rorschacherstrasse 95, 9011 St. Gallen, Switzerland.

## Abstract

**Background:** There is insufficient evidence regarding the role of respirators in the prevention of SARS-CoV-2 infection. We analysed the impact of filtering facepiece class 2 (FFP2) *vs*. surgical masks on the risk of SARS-CoV-2 acquisition among Swiss healthcare workers (HCW).

**Methods:** Our prospective multicentre cohort enrolled patient-facing HCWs from June to August 2020. Participants were asked about COVID-19 risk exposures/behaviours, including preferred mask type when caring for COVID-19 patients outside of aerosol-generating procedures (AGP). For those performing AGPs, we asked whether they used FFP2 irrespective of the patient’s COVID-19 status (i.e. universal use). The impact of FFP2 on i) self-reported SARS-CoV-2-positive nasopharyngeal PCR/rapid antigen tests captured during weekly surveys, and ii) SARS-CoV-2 seroconversion between baseline and January/February 2021 was assessed.

**Results:** We enrolled 3’259 participants from nine healthcare institutions, whereof 716 (22%) preferentially used FFP2 respirators. Among these, 81/716 (11%) reported a SARS-CoV-2-positive swab, compared to 352/2543 (14%) surgical mask users (median follow-up 242 days); seroconversion was documented in 85/656 (13%) FFP2 and 426/2255 (19%) surgical mask users. Adjusted for baseline characteristics, COVID-19 exposure, and risk behaviour, FFP2 use was non-significantly associated with a decreased risk for SARS-CoV-2-positive swab (adjusted hazard ratio [aHR] 0·8, 95% CI 0·6-1·0, p=0·052) and seroconversion (adjusted odds ratio [aOR] 0·7, 95% CI 0·5-1·0, p=0·053); household exposure was the strongest risk factor (aHR for positive swab 10·1, p<0·001; aOR for seroconversion 5·0, p<0·001). In subgroup analysis, FFP2 use was clearly protective among those with frequent (>20 patients) COVID-19 exposure (aHR 0·7, p<0·001; aOR 0·6, p=0·035). Universal FFP2 use during AGPs showed no protective effect (aHR 1·1, p=0·7; aOR 0·9, p=0·53).

**Conclusion:** Respirators compared to surgical masks may convey additional protection from SARS-CoV-2 for HCW with frequent exposure to COVID-19 patients.

**Funding:** Swiss National Sciences Foundation, Federal Office of Public Health, Cantonal Health Department St.Gallen

## INTRODUCTION

The most common mode of Severe Acute Respiratory Syndrome Coronavirus 2 (SARS-CoV-2) transmission is believed to be via respiratory droplets and – probably less important – via fomites.^1,2^ The role of microdroplets or aerosols, which might travel beyond one to two meters from an infected person, as a potential mode of SARS-CoV-2 transmission has been heavily debated.^3–6^ Reports from healthcare and non-healthcare settings suggest that SARS-CoV-2 may indeed be transmitted via aerosols, particularly in poorly ventilated indoor environments, even outside of so-called aerosol-generating procedures (AGPs).^7,8^

Healthcare workers (HCW) remain at the front line, with a high risk of exposure to and infection with SARS-CoV-2.^9^ For HCW involved in AGPs, international guidelines unanimously recommend the use of so-called respirators, which include filtering facepiece class 2 (FFP2), N95, or KN95, and which have the ability to filter microparticles. In contrast to respirators, surgical (also termed medical) masks are designed to provide barrier protection and only block larger respiratory droplets. As a consequence of the conflicting opinions about aerosol transmission, guidelines differ regarding recommendations for the use of respirators outside of AGPs. Whereas the Centers of Disease Control and Prevention (CDC) and the European Centre for Disease Prevention and Control (ECDC) recommend respirators if available, the World Health Organization (WHO) and the Swiss National Centre for Infection Prevention (Swissnoso) recommend surgical masks.^10–13^ The Infectious Diseases Society of America’s recommendation is to use either a surgical mask or a respirator.^14^

Prospective head-to-head comparisons evaluating the protective effect of these mask types against SARS-CoV-2 acquisition are sparse. In a meta-analysis including mostly studies on non-SARS-CoV-2 coronaviruses, use of respirators was associated with a stronger protective effect, although no study directly compared respirators to surgical masks.^15^ A meta-analysis comparing the clinical effectiveness of respirators to surgical masks for other respiratory viruses, including coronaviruses, found no significant difference concerning infection risk in HCW.^16^

For SARS-CoV-2, we identified three studies not covered in the above-mentioned meta-analysis. An online survey among HCW from different countries showed a protective effect of respirators compared to surgical masks for those performing AGPs on Coronavirus Disease 2019 (COVID-19) patients.^17^ A cross-sectional study from the US (in preprint) found respirator use to be associated with decreased seropositivity rate, although no multivariate analysis was perforned.^18^ In a prospective single-centre HCW cohort from Western Switzerland, respirators were protective regarding SARS-CoV-2 seroconversion, although use of other PPE was not documented and residual confounding was suspected.^19^

To summarize, there is currently insufficient evidence to determine if the use of FFP2 respirators reduces the risk of SARS-CoV-2 infections. In this analysis of prospective cohort data from Swiss HCWs, we sought to assess the effectiveness of FFP2 compared to surgical masks regarding SARS-CoV-2 protection for HCW involved in patient care.

## METHOD

### Study design, participants and setting

We performed a prospective observational multicentre cohort consisting of employees (aged 16 years or older) from different healthcare institutions in Northern and Eastern Switzerland (from four different cantons). All hospital employees irrespective of involvement in patient care were eligible to participate; however, for the current analysis we excluded those without patient contact. Employees registered online and provided electronic consent. Enrolment took place from June 22^nd^ to August 15^th^ 2020, between the end of the first COVID-19 wave in Switzerland and the surge of the second wave;^20^ data were analysed up to March 9^th^ 2021, when the second wave had abated (**Figure 1**). The study was approved by the ethics committee of Eastern Switzerland (#2020-00502).

**Figure 1.**
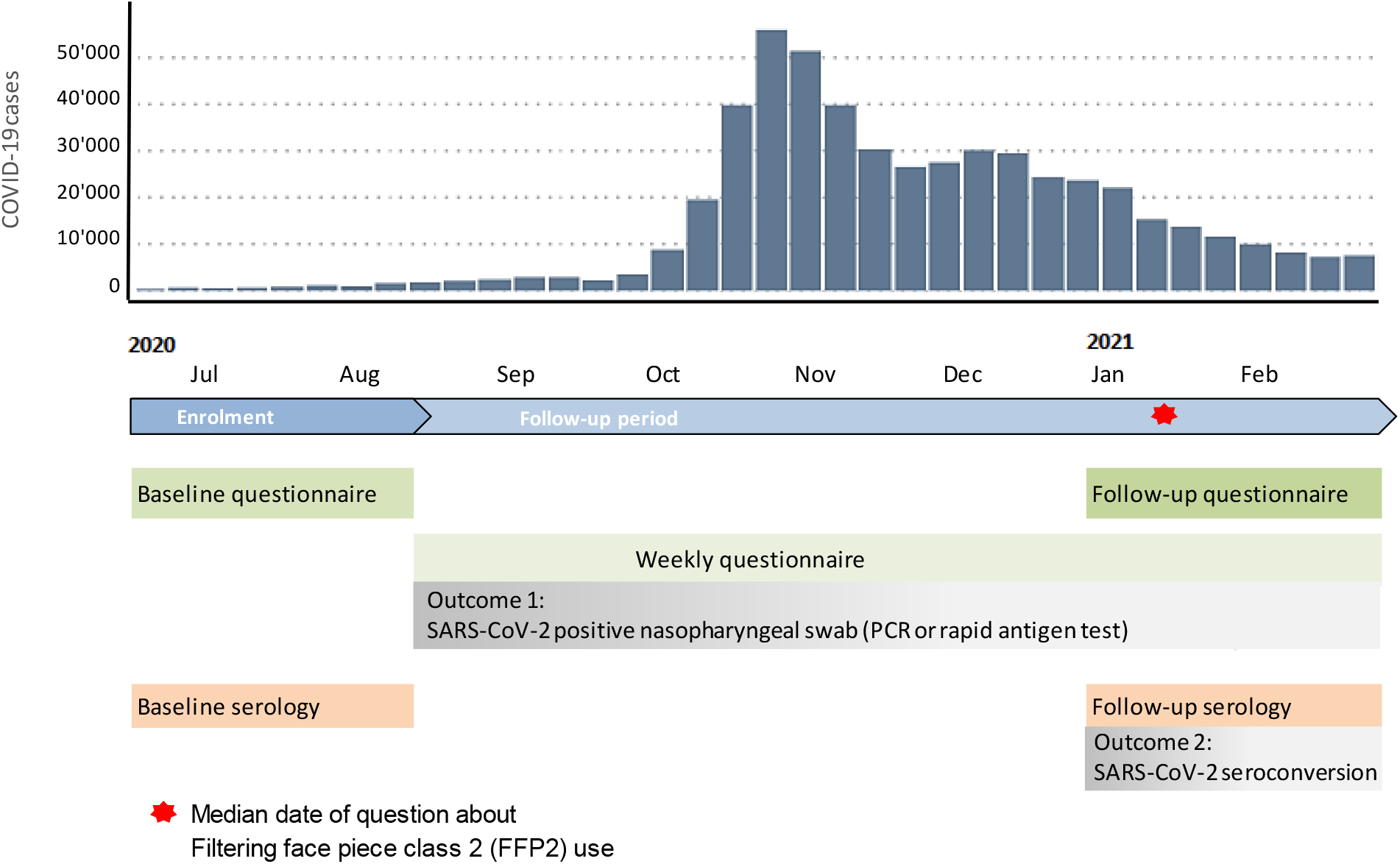
Study timeline and procedures in relation to laboratory confirmed (by polymerase chain reaction [PCR] or rapid antigen test) COVID-19 cases in Switzerland (absolute number of weekly cases).^31^

### National and local mask policies

During the study period, a national policy required Swiss residents (including HCW) to wear at least a surgical mask at work. The Swiss National Centre for Infection Prevention (Swissnoso) suggests the use of a respirator mask only while performing AGPs on confirmed or suspected COVID-19 patients.^13^ However, this was considered minimum standard and institutions were free to recommend respirators outside of AGPs. Also, in most institutions HCW could make the personal choice of wearing a respirator or surgical mask at work. To characterize institution-level mask recommendation, we conducted a survey of representatives from participating institutions asking about local policies for FFP2 use and estimated compliance with those policies (among a choice of “good”, “more FFP2 used than recommended”, “less FFP2 used than recommended)”.

### Study procedures and questionnaires

The study timeline is shown in **Figure 1**. Upon inclusion, participants answered a baseline questionnaire asking about anthropometric data, pregnancy and comorbidities, job description (including full-time equivalent [FTE] percentage, profession, involvement in AGPs, working in intensive care, exposure to COVID-19 patients, use of PPE, or visit to staff restaurant), and non-work related risk behaviour such as wearing a mask outside of work, leisure and shopping activities, but also opinions on adequacy of regulatory measures. During follow-up, participants received weekly text messages and emails with an electronic link to a questionnaire where they indicated results of nasopharyngeal swabs (polymerase chain reaction or rapid antigen tests) for SARS-CoV-2. Additionally, participants answered whether they had been exposed to confirmed COVID-19 patients, co-workers, household contacts, or other COVID-19 cases during the previous week. In January 2021, (i.e. before follow-up serology was performed), a follow-up questionnaire asked about use of mask type (FFP2 *vs*. surgical mask) *outside* of AGPs. For those involved in care of COVID-19 patients, this question explicitely asked about mask use during COVID-19 patient contact. AGPs were defined as bronchoscopies, in-/extubation, gastroscopy, transesophageal echocardiography, reanimation, non-invasive ventilation, and suction of tracheal secretions. Participants had the choice among “Use of surgical mask only”; “Mostly use of surgical mask”; “Equal use of both mask types”; “Mostly use of FFP2”; “Use of FFP2 only”. For the purpose of this analysis, the two latter categories were classified as “Mostly FFP2”, whereas the first three categories were merged into the group “Not mostly FFP2” (for better comprehensibility termed “Mostly surgical masks”). For HCW involved in AGPs, we also asked whether they always used FFP2 during AGPs, irrespective of the patient’s COVID-19 status (i.e. termed “universal FFP2 use”). Furthermore, use of other PPE including gowns, gloves and goggles while caring for COVID-19 patients was asked, as well as the number of COVID-19 patients and of positive co-workers HCW had been knowingly exposed since March 2020 (for description and categorisation of variables, see **Table S1**).

### Outcome assessment

Two main outcomes were defined: i) time to the first self-reported SARS-CoV-2 positive nasopharyngeal swab and ii) SARS-CoV-2 seroconversion. Results of nasopharyngeal swabs were asked in the weekly questionnaires. Baseline (June – August 2020) and follow-up (January/February 2021) serologies were performed to assess SARS-CoV-2 seroconversion (**Figure 1**). Participants with positive serology at baseline were excluded from this analysis. Samples were analysed with an electro-chemiluminescence immunoassay (ECLIA, Roche Diagnostics, Rotkreuz, Switzerland, detection of total antibodies directed against the nucleocapsid-(N)-protein of SARS-CoV-2), as described elsewhere.^20^

### Statistical analysis

For analysis of nasopharyngeal swabs, we performed Cox regression with time intervals between consecutive weekly questionnaires as response. Intervals were censored as long as no positive swab (event) was reported, and those following the first event were excluded. The model included COVID-19 exposures reported in any of the three weekly questionnaires submitted before the end of each time interval as well as the cumulative number of negative swabs up to an interval’s end (to account for different testing behaviour) as time-dependent co-variables, and answers from the baseline and follow-up questionnaire (including mask type) as time-independent co-variables. These variables were *a priori* chosen from the baseline questionnaire,^20^ based on their expected potential to confound the association between mask use and risk for SARS-CoV-2 acquisition. Cantons and institutions were included as cluster terms. For seroconversion, we used logistic regression including the same time-independent co-variables as for nasopharyngeal swabs as fixed effects and cluster terms as random effects. Instead of the time-dependent covariables, we included overall household exposure (summarizing weekly reports into “any” *vs*. “none”) as well as the total number of COVID-19 patient and co-worker exposures. Variance inflation factors (VIF) were calculated to check that no co-variable had a VIF>5. R statistical software Version 4.0.2 was used for all statistical analyses.

### Sensitivity and subgroup analyses

We performed three sensitivity analyses to assess the influence of possible confounders on the estimated effect of mask type on the two outcomes: (1) excluding HCW with COVID-19 cases within their household (which might reduce the importance of professional exposure and protection), (2) including cantons and institution as fixed effects (to account for regional incidence and institutional factors), and (3) excluding HCW tested positively before December 1^st^ 2020 (because mask use may have changed through time and was asked only in January 2021).

Because mask type was likely to be most important for HCW with frequent COVID-19 exposure, we performed a subgroup analysis according to frequency of COVID-19 patient contact (no known contact *vs*. 1-20 patients *vs*. >20 patients since March 2020). Also, we repeated the analysis for those performing AGPs, using a model including whether they always used FFP2 during AGPs.

## RESULTS

### Institutions

We included participants from seven acute care institutions (with 14 different sites), one rehabiliation clinic, and three psychiatry clinics (analysed as one institution). Policies on respiratory protection according to institution are summarized in **Table 1**. Most institutions followed the Swissnoso recommendations for use of FFP2 respirators. Actual FFP2 use varied considerably between institutions and ranged from 3% to 52% of participants. All acute care institutions with less strict local guidelines (i.e. no FFP2 mask required during contact with COVID-19 patients) reported that FFP2 masks were more frequently used than recommended (**Table 1**).

**Table 1.** List of participating hospitals with local mask policies (FFP2 or surgical masks), self-reported global adherence, FFP2 use based on study data, and SARS-CoV-2 seroprevalence among healthcare workers.

| Clinic | AGPs |  | Non-AGPs |  | Hospital area | Fit test | Adherence | HCW (n) | Reported FFP2 use (in %) | Sero-prevalence (in %) |
| --- | --- | --- | --- | --- | --- | --- | --- | --- | --- | --- |
|  | COVID-19 patients | Other patients | COVID-19 patients | Other patients |  |  |  |  |  |  |
| A | FFP2 | Surgical | FFP2 | Surgical | Surgical | Yes | Good | 612 | 34.6 | 10.8 |
| B | FFP2 | Surgical | Surgical | Surgical | Surgical | No | More FFP2 | 241 | 51.5 | 17.0 |
| C | FFP2 | Surgical | Surgical | Surgical | Surgical | No | More FFP2 | 105 | 13.3 | 37.1 |
| D | FFP2 | Surgical | Surgical | Surgical | Surgical | No | More FFP2 | 329 | 12.2 | 30.4 |
| E<sup>1</sup> | FFP2 | Surgical | Surgical | Surgical | Surgical | No | More FFP2 | 882 | 10.5 | 20.1 |
| F | FFP2 | Surgical | Surgical | Surgical | Surgical | No | More FFP2 | 94 | 16.0 | 14.9 |
| G | FFP2 | FFP2 | FFP2 | Surgical | Surgical | No | Good | 299 | 43.8 | 11.7 |
| H<sup>2</sup> | NA | NA | Surgical | Surgical | Surgical | No | Good | 262 | 9.2 | 11.1 |
| I<sup>3</sup> | FFP2 | FFP2 | Surgical | Surgical | Surgical | No | Good | 87 | 3.4 | 11.5 |
| Swiss-noso<sup>4</sup> | FFP2 | Surgical | Surgical | Surgical | .. | .. | .. | .. | .. | .. |

<sup>1</sup> Includes geriatric hospital
<sup>2</sup> Three psychiatry clinics
<sup>3</sup> Rehabilitation clinic
<sup>4</sup> National recommendations<sup>13</sup>
Abbreviations: FFP2 – Filtering facepiece class 2. AGP – Aerosol-generating procedure. HCW – Health care worker. Surgical – Surgical mask. NA – Not applicable.

### Baseline characteristics

Among the 3’259 participants, 614 (19%) were male and median age was 39 years (interquartile range [IQR] 30-49 years). Most were nurses (n=1’724, 53%), followed by physicians (n=671, 21%). Preferential use of FFP2 while caring for COVID-19 patients was reported by 716 (22%). HCW who preferred using FFP2 were more likely to be male (OR 1·5, p<0·001), to be ≤ 50 years old (OR 1·4, p=0·001), to support stronger public restrictions regarding the pandemic (OR 1.7, p<0·001), to be involved in AGPs (OR 4·2, p<0·001), to work in intensive care (OR 8·4, p<0·001), to be exposed to >20 COVID-19 patients (OR 2.8, p<0·001), to use gowns (OR 8·7, p<0.001), gloves (OR 2·5, p<0.001) and goggles (OR 8·0, p<0.001) while caring for COVID-19 patients, and to undergo testing for SARS-CoV-2 (OR 1·4, p<0·001) (**Table 2**).

**Table 2.** Factors associated with preferential use of FFP2 *vs*. surgical masks among 3’259 healthcare workers.

| Risk or protection factor for SARS-CoV-2 infection | Number of HCW without and with the factor |  | Frequency of the factor (n and % with it) in relation to FFP2 use during contact with COVID-19 patients |  | OR (95% CI) | p-value (Fisher's test) |
| --- | --- | --- | --- | --- | --- | --- |
|  | without | with | Mostly surgical mask | Mostly FFP2 |  |  |
|  |  |  | (n = 2543) | (n = 716) |  |  |
| **Sociodemographic data** |  |  |  |  |  |  |
| Age > 50 years | 2528 | 731 | 604 (24%) | 127 (18%) | 0.69 (0.56 - 0.86) | 0.001 |
| Sex: male | 2645 | 614 | 444 (17%) | 170 (24%) | 1.47 (1.20 - 1.81) | < 0.001 |
| Living in Germany or Austria | 3094 | 165 | 104 (4%) | 61 (9%) | 2.18 (1.55 - 3.06) | < 0.001 |
| Child in household | 2424 | 835 | 650 (26%) | 185 (26%) | 1.01 (0.83 - 1.23) | 0.884 |
| **Medical conditions** |  |  |  |  |  |  |
| Comorbidity | 2066 | 1193 | 921 (36%) | 272 (38%) | 1.08 (0.91 - 1.28) | 0.404 |
| Active smoker | 2719 | 540 | 426 (17%) | 114 (16%) | 0.94 (0.74 - 1.18) | 0.649 |
| Pregnant during study | 3122 | 137 | 113 (4%) | 24 (3%) | 0.75 (0.46 - 1.18) | 0.246 |
| **Behaviour outside of work** |  |  |  |  |  |  |
| Prophylactic home remedies | 2787 | 472 | 363 (14%) | 109 (15%) | 1.08 (0.85 - 1.37) | 0.548 |
| Social leisure activities | 1693 | 1566 | 1208 (48%) | 358 (50%) | 1.11 (0.93 - 1.31) | 0.253 |
| Wearing a mask outside work | 2354 | 905 | 657 (26%) | 248 (35%) | 1.52 (1.27 - 1.82) | < 0.001 |
| Support for stronger public restrictions | 2643 | 616 | 433 (17%) | 183 (26%) | 1.67 (1.37 - 2.05) | < 0.001 |
| **HCW specifics** |  |  |  |  |  |  |
| Job: Nurse | 1535 | 1724 | 1309 (51%) | 415 (58%) | 1.30 (1.10 - 1.54) | 0.002 |
| Job: Physician | 2588 | 671 | 500 (20%) | 171 (24%) | 1.28 (1.05 - 1.57) | 0.016 |
| Full-time job (> 80%) | 1488 | 1771 | 1325 (52%) | 446 (62%) | 1.52 (1.28 - 1.81) | < 0.001 |
| Involved in AGP | 2055 | 1204 | 747 (29%) | 457 (64%) | 4.24 (3.55 - 5.07) | < 0.001 |
| Work in intensive care | 2971 | 288 | 102 (4%) | 186 (26%) | 8.39 (6.43 - 10.99) | < 0.001 |
| Contact to >20 COVID-19 patients | 1812 | 1120 | 732 (32%) | 388 (58%) | 2.83 (2.36 - 3.39) | < 0.001 |
| **Behaviour at work** |  |  |  |  |  |  |
| Hygiene knowledge | 491 | 2768 | 2121 (83%) | 647 (90%) | 1.87 (1.42 - 2.48) | < 0.001 |
| Regular meals in staff restaurant | 1094 | 2165 | 1702 (67%) | 463 (65%) | 0.90 (0.76 - 1.08) | 0.263 |
| Handwashing more frequent | 343 | 2916 | 2268 (89%) | 648 (91%) | 1.16 (0.87 - 1.55) | 0.335 |
| **Test results** |  |  |  |  |  |  |
| Positive SARS-CoV-2 test<sup>1</sup> | 2826 | 433 | 352 (14%) | 81 (11%) | 0.79 (0.61 - 1.03) | 0.081 |
| Negative SARS-CoV-2 test<sup>1</sup> | 1633 | 1626 | 1231 (48%) | 395 (55%) | 1.31 (1.11 - 1.55) | < 0.001 |
| **Use of PPE** |  |  |  |  |  |  |
| Always used goggles<sup>2</sup> | 2362 | 897 | 446 (18%) | 451 (63%) | 7.99 (6.64 - 9.65) | < 0.001 |
| Always used gloves<sup>2</sup> | 1939 | 1320 | 904 (36%) | 416 (58%) | 2.51 (2.12 - 2.99) | < 0.001 |
| Always used a gown<sup>2</sup> | 2362 | 897 | 437 (17%) | 460 (64%) | 8.65 (7.17 - 10.46) | < 0.001 |
| Always used FFP2 during AGP | 645 | 559 | 242 (32%) | 317 (69%) | 4.72 (3.65 - 6.12) | < 0.001 |

<sup>1</sup>at least one positive/negative test. The mean number of negative tests per HCW was 0.90 for surgical mask users and 1.11 for FFP2 users.
<sup>2</sup>in contact with COVID-19 patients outside of AGPs
Abbreviations: FFP2 – Filtering facepiece class 2. HCW – Health care worker. AGP – Aerosol-generating procedure. PPE – Personal protective equipment. NA – Not applicable.

### Risk of positive SARS-CoV-2 test according to mask type

Median follow-up was 242 days, both for respirator and for surgical mask users (Wilcoxon test, p=0·49). The number of self-reported positive SARS-CoV-2 tests was 81/716 (11%) for FFP2 users compared to 352/2543 (14%) in users of surgical masks (hazard ratio [HR] 0·8; 95% confidence interval [CI] 0·6-1·0; p=0·06 log-rank test). In the Cox regression model, the factor most strongly associated with a positive SARS-CoV-2 test was exposure to a positive household contact (adjusted HR [aHR] 10·1, 95% CI 7·5-13·5, p<0·001). Use of FFP2 while caring for COVID-19 patients was associated with a decreased risk of acquiring SARS-CoV-2 (aHR 0·8, 95% CI 0·7-1·0, P=0·052) (**Figure 2, Table S2**). In sensitivity analyses, restriction to data collected after December 1^st^ (aHR 0·7, p=0.03) showed similar results; removing participants with a positive household member (aHR 0·8, p=0.31) and treating institutions/cantons as fixed effect (aHR 0·9, p=0.43) resulted in non-significant associations (**Table S2**).

**Figure 2.**
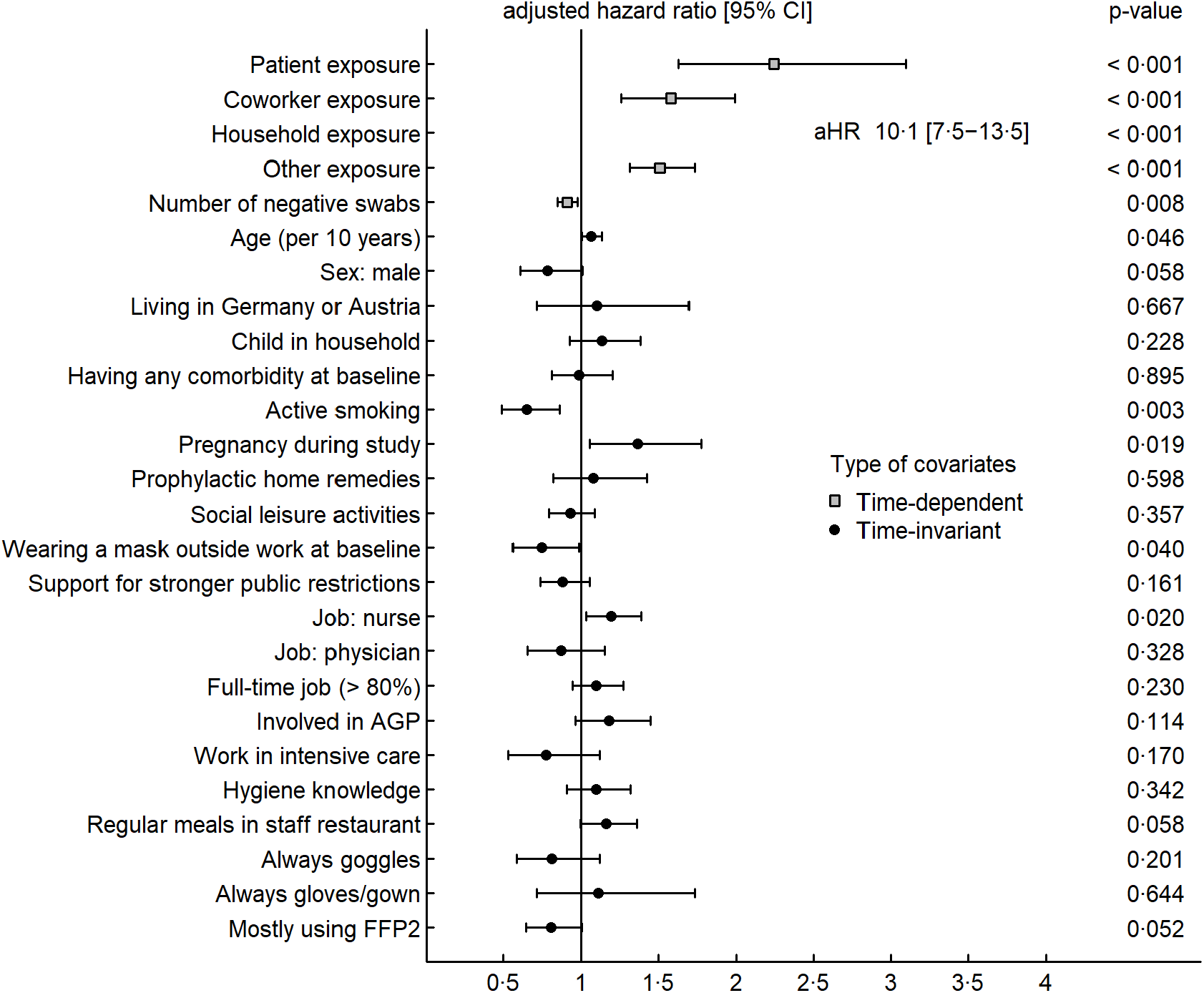
Cox regression analysis with outcome “SARS-CoV-2- positive nasopharyngeal PCR/rapid antigen test” (participants n=3’259, positive swabs n=433). Abbreviations: AGP – Aerosol-generating procedure. FFP2 – Filtering facepiece class 2.

### Risk of SARS-CoV-2 seroconversion according to mask type

We included 2’916 HCW with negative baseline serology, who had a second serology performed in January/February 2021. Seroprevalence was 12·9% (85/658) for FFP2 users compared to 18·9% (429/2258) for users of surgical masks (OR 0·6, 95% CI 0·5-0·8, p<0·001). In multivariable analysis, the strongest risk factor for seroconversion was again having a positive household member with an adjusted odds ratio (aOR) of 5·0 (95% CI 3·9-6·5, p<0·001). FFP2 use was non-significantly associated with decreased risk for seroconversion (0·7, 95% CI 0·5-1·0, p=0.053) (**Figure 3, Table S3**). In sensitivity analyses, removal of variable on household exposure (aOR 0·7, p=0.046) and including cantons/institutions as fixed effects (aOR 0·8, p=0.088) did not significantly change the point estimates for FFP2 use nor the significance levels (**Table S3**).

**Figure 3.**
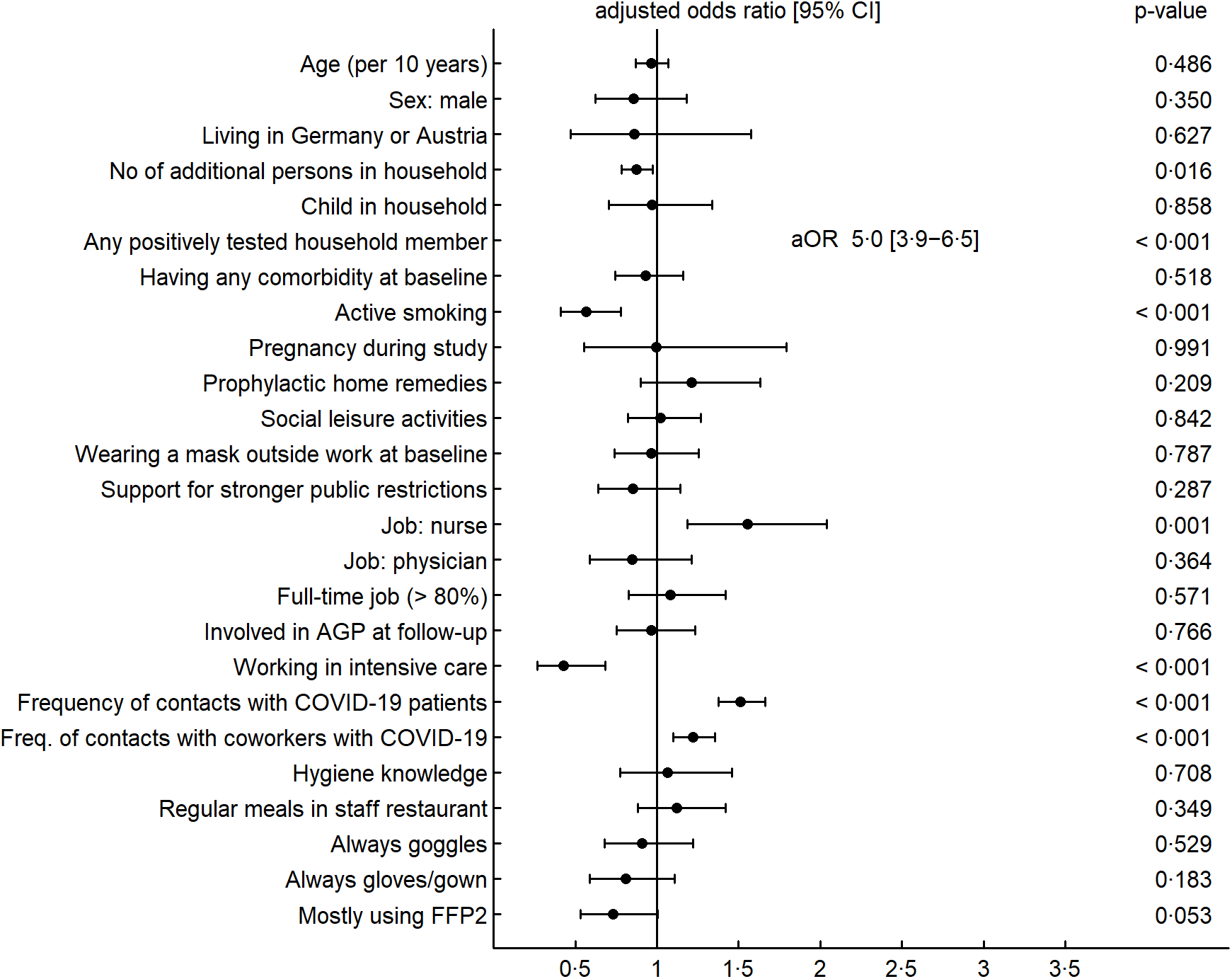
Forest plots showing results of the generalized mixed-effects model regarding outcome “SARS-CoV-2 seroconversion” (participants n=2’916, seroconversions n=511). Abbreviations: AGP – Aerosol-generating procedure. FFP2 – Filtering facepiece class 2.

### Subgroup analyses

HCW with frequent (>20) exposure to COVID-19 patients were less likely to report a positive SARS-CoV-2 swab when mostly wearing FFP2, with an unadjusted HR of 0·6 (p<0·001) compared to 0·8 (p=0·18) for those exposed to 1-20 patients (**Figure 4**). Results of multivariable analyses were similar to those in univariable analysis, both for positive swabs (aHR 0·7, p<0·001, *vs*. aHR 1·1, p=0·77) and seroconversion (aOR 0·6, p=0·036, *vs*. aOR 0·8, p=0·32) (**Table S4**). The number of positive swabs for HCW without COVID-19 patient contact was too small to perform multivariable analyses (2 events among 40 FFP2 users, 29 events among 480 surgical mask users).

**Figure 4.**
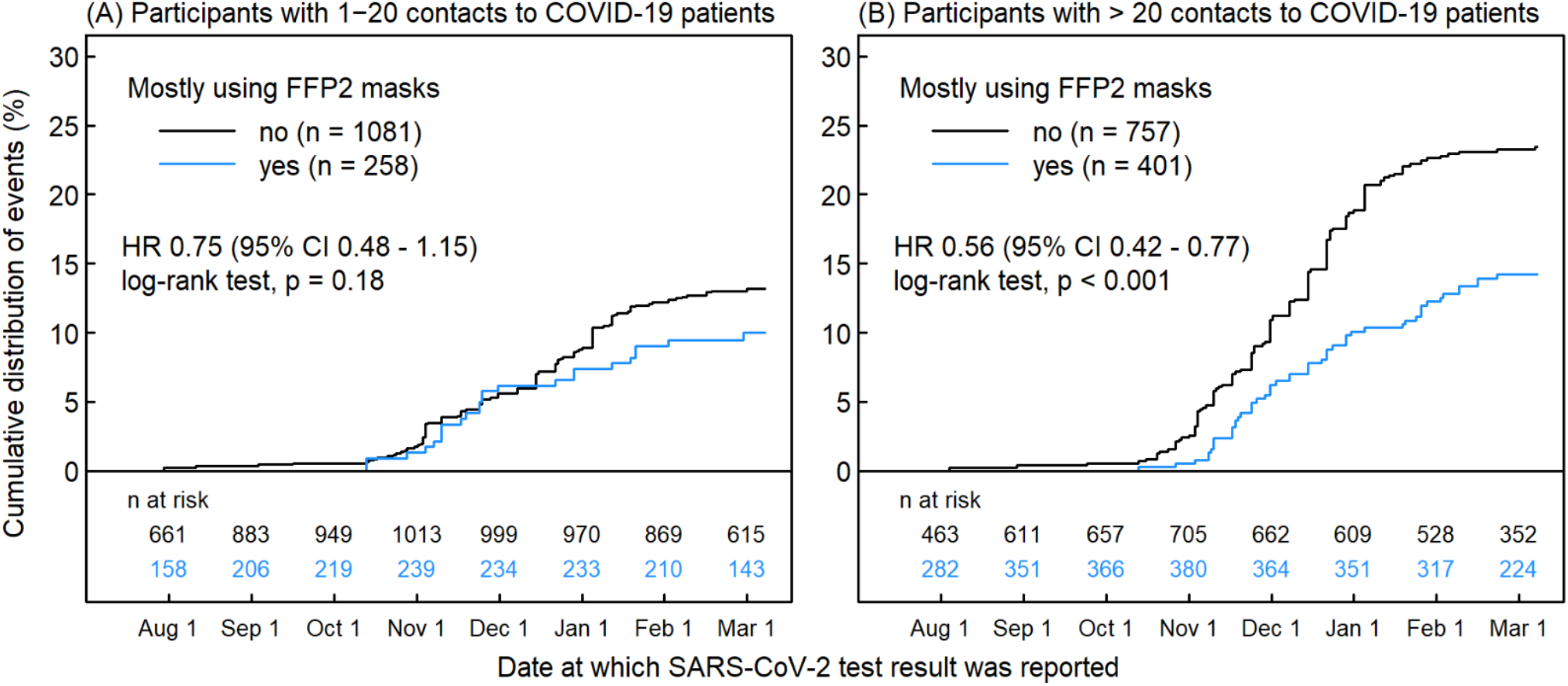
Kaplan-Meier curve regarding unadjusted risk of a SARS-CoV-2-positive nasopharyngeal swab for HCW with infrequent (A) and frequent (B) exposure to COVID-19 patients. Abbreviations: HCW – Health care worker. FFP2 – Filtering facepiece class 2.

For HCW performing AGPs, universal use of FFP2 during AGPs (irrespective of the patients COVID-19 status) showed no effect (aHR 1·1, p=0·66 and aOR 0·9, p=0·54, respectively) (**Table S5**).

## DISCUSSION

In this prospective multicentre HCW cohort, FFP2 use outside of AGPs was marginally associated with a decreased risk for SARS-CoV-2 infection when compared to surgical masks. The effect was however pronounced for those with frequent COVID-19 patient exposure. Using FFP2 irrespective of the patient’s COVID-19 status did not provide additional protection for HCWs involved in AGPs. The large sample size, the dual approach for outcome assessment, and consideration of a variety of potential confounder variables (including personal risk factors, use of other PPE, and general risk perception) are among the strengths of this study.

This is, to our knowledge, the first prospective multicentre study comparing the effect of respirators and surgical masks regarding protection from SARS-CoV-2. In Switzerland, national guidelines do not recommend the use of respirators for HCW caring for COVID-19 patients outside of AGPs; at the same time institutional guidelines are heterogeneous. This unique setting allowed us to study this important research question. The overall association between FFP2 use and risk for SARS-CoV-2 infection was just marginally non-significant. This and the inconsistencies seen in our sensitivity analyses are probably a reflection of the heterogeneous study population, which consisted by two thirds of HCWs with only sporadic (or even no known) COVID-19 exposure.

However, for HCW with more frequent exposure, we found a clearly significant protective effect associated with FFP2 use. Several reports suggest that aerosol transmission is indeed a non-negligible mode of SARS-CoV-2 transmission and that respirator masks may provide additional protection compared to surgical masks.^17–19,21,22^ On the other hand, single case reports have suggested that surgical masks are equivalent to respirators in protecting HCW from SARS-CoV-2 infection.^23,24^ These supposedly contradictory findings can be reconciled when considering a particular feature of SARS-CoV-2, namely its high overdispersion.^25^ Overdispersion describes the highly variable transmissibility of infected individuals; in other words, only a minority of infected individuals actually transmit the virus to others, often within so-called superspreading events. As a consequence, the probability of being exposed but not infected – as seen in the case reports above – is relatively high (irrespective of mask type). Supporting this hypothesis, a simulation study by Chen *et al*. describes how the SARS-CoV-2 viral load of infectious individuals can vary largely and how this influences infection probability and the effectiveness of the different mask types.^26^

Notably, we did not observe any protective effect of FFP2 for HCW performing AGPs in the absence of any COVID-19 suspicion in the patient. We extrapolate from these findings that universal FFP2 use in the hospital setting, where the average risk exposure is usually lower than during AGPs, does not provide additional protection compared to surgical masks. We acknowledge however that in settings with a high proportion of undiagnosed, asymptomatic or presymptomatic patients, an additional benefit through universal FFP2 use cannot be excluded. Our effect size (aOR 0.7) was in the range of those reported by Lentz *et al*. (aOR of 0.4) and Martischang *et al*. (aOR of 0.7).^17,19^ Given the fact that many HCWs in our study did not consistently wear either FFP2 or surgical masks, we assume that the protective effect of FFP2 might be even higher in reality. However, the clinical significance of the protective effect mediated by FFP2 use can be questioned, given the dominating impact of extra-occupational risk exposures on the COVID-19 risk of HCWs, as seen in other studies.^27^ Also, the disadvantages of respirators – the discomfort of wearing such masks over long periods of time, the fact that their protective effect can be diminished without prior HCW training and fit testing (which was indeed not done in most of our institutions), as well as their cost – have to be considered when assessing the net benefit of FFP2 over surgical masks.^28,29^

To adjust for potential confounding, we included the use of gloves, gowns and goggles in our multivariable analysis. None of these measures were associated with any clear additional protective effect. Further studies, ideally randomized controlled trials, are needed to evaluate the role of these components of PPE in SARS-CoV-2 protection. Other associations with SARS-CoV-2 infection found in our study, such as the “protective” effect of active smoking or the increased risk associated with working as a nurse, have been discussed earlier^20^.

Our study has limitations. First, residual confounding is possible. Yet, we have included multiple co-variables accounting for risk exposures and risk behaviours. Also, the fact that use of other PPE or universal respirator use among HCW performing AGPs (representing HCW with particularly risk-averse behaviour) were not associated with reduced seroconversion rate, supports our argument of a valid multivariable model with low risk of residual confounding. Second, the question about respirator use was asked in a questionnaire in January 2021, at a time when most SARS-CoV-2 positive participants had already had their infection. A positive test result could have led to a change in preferred mask type (in either direction). However, restricting the analysis to the time period close to the follow-up questionnaire showed similar or even stronger assocations compared to the full model. Third, we did not specifically ask about duration of contact to individual COVID-19 patients, although type of profession, work percentage, or involvement in AGP can be regarded as proxy for this potentially important variable. Fourth, although we included multiple institutions, settings, and geographical regions in our study, the generalizability of the results can be questioned due to the fact that participation in the study was non-mandatory. However, distribution of key variables (e.g. age, sex, profession) were similar between the total HCW population (from the largest participating institution) and the cohort population.^20^

To conclude, FFP2 use outside of AGPs is associated with a reduced risk of SARS-CoV-2 acquisition for HCW with high COVID-19 exposure, while those with only sporadic or no known contact do not seem to benefit. The effect size should be interpreted in the context of the global COVID-19 risk for HCW, which is driven by exposure to positive household contacts. No significant protective effect was observed for those using FFP2 during AGPs in the absence of clinical COVID-19 suspicion. Pending results of randomized controlled trials,^30^ our data are an important step for healthcare institutions and policy makers to gauge the expected add-on value of respirators compared to surgical masks.

## Data Availability

All data are available from the corresponding author upon reasonable request.

## Author contributions

SG, MS, PV, CRK and PK conceptualized the study; SK, AM, PV, CRK and PK were involved in funding acquisition; TE, OL, DF, AB, EL, JCM, PR, MR, RS, DV, BW, UB, CRK and PK were involved in data collection; TE and LR were responsible for laboratory analyses; SH, SG, TE, CRK and PK were responsible for data analysis; SH, GS and RT performed the literature search; SH and SG were responsible for creation of figures; SH, SG, TE, MS, CRK and PK interpreted the data; CRK and PK supervised the study.

