## Supplements for "Use of respirator vs. surgical masks in healthcare personnel and its impact on SARS-CoV-2 acquisition – a prospective multicentre cohort study"

Table S1. Description and categorization of variables used in analyses.

| Variable | Question in Questionnaire | Answer possibilities | Calculation / Categorisation | Timepoint |
|---|---|---|---|---|
| **Test results** | | | | |
| Positive SARS-CoV-2 test | Did you have a positive SARS CoV-2 swab test? | True; False | True; False | Weekly |
| Number of negative swab tests | Have you been tested for SARS-CoV-2 in the past week? | True; False | Cumulative number of negative tests up to this date | Weekly |
| | What was the result? | Positive; Negative; Pending | | Weekly |
| **Exposure to COVID-19** | | | | |
| Frequency of contacts to COVID-19 patients | How many COVID-19 patients were you exposed to since March 2020? | None; 1 to 5; 6 to 10; 11 to 20; more than 20; I don't know | None, 1-20, >20 | Follow-up |
| Frequency of contacts to coworkers with COVID-19 | How many SARS-CoV-2 confirmed coworkers were you exposed to since March 2020? | 1; 2; 3; 4 or more | 1;2;3;4 | Follow-up |
| Any positively tested household member | Has someone of your household or your intimate partner been tested positive since March 2020? | True; False | True; False | Weekly |
| Patient exposure *(time dependent)* | Did you have contact to a confirmed COVID-19 patient in hospital last week? | True; False | True; False | Weekly |
| Coworker exposure *(time dependent)* | Did you have contact to a SARS-CoV-2 confirmed coworker whithout wearing a mask last week? | True; False | True; False | Weekly |
| Household exposure *(time dependent)* | Has someone from your household been tested positive for SARS-CoV-2 last week? | True; False | True; False | Weekly |
| Other exposure *(time dependent)* | Has someone other from your surroundings been tested positive for SARS-CoV-2 last week? | True; False | True; False | Weekly |
| **Sociodemographic data** | | | | |
| Age | What is your date of birth? | Day/Month/Year | (2020−birthyear)/10 | Baseline |
| Sex | What is your sex? | Male; Female; Not specified | Male; Female and other | Baseline |
| Living in Germany or Austria | Zip code of your hometown | Numbers | GE/AU *vs.* CH | Baseline |
| Number of additional persons in household | How many people live in your place of residence? | Numbers | Numbers from 0 to 4 (larger numbers truncated to 4) | Baseline |
| Child in household | Year of birth for every household contact | Year | At least one year ≥ 2008 | Baseline |
| **Medical conditions** | | | | |
| Comorbidity | Do you suffer from any of the following diseases? | None; Arterial hypertension; Diabetes mellitus; Heart disease; COPD; Asthma; Liver disease (Fatty liver; Cirrhosis); Cancer; Rheumatologic disease (Arthritis, Lupus, Vasculitis); Hay fever; Hypothyroidism; Chronic inflamatory bowel disease (M. Crohn, Colitis ulcerosa), Intermittent claudication (Peripheral artery disease), Stroke | None = False; All others = True | Baseline |
| Active smoker | Do you smoke or did you use to smoke? | No; Used to smoke; Active smoker | Active smoker; other | Baseline |
| Pregnancy during study | Are you pregnant at the moment? | True; False | True; False | Follow-up |
| **Behaviour outside of work** | | | | |
| Prophylactic home remedies | Which preventive preparation do you take against COVID-19? | None; Echinacea; Zinc; Vitamin C; Umckaloabo/Kaloba; Vitamin B12; | None = False; All others = | Baseline |

| | | | | |
|---|---|---|---|---|
| | | Homeopathics; Others | True | |
| Social leisure activities | Which activities do you perform regularly at the moment? | None; Sportsclub; Choir; Orchestra/Music group (Active musician); Concert/Theatre; Cinema; Church; Yoga/Fitness; Restaurant/Bar | None = False; All others = True | Baseline |
| Wearing a mask outside work | In which situations did you wear a mask? | At home; Outside; On public transport; While shopping; In hospital area; In the workingspace; During contact with other people | None = False; All others = True | Baseline |
| Support for stronger public restrictions | Do you support the national measures and it's duration or what is your opinion about it? | 1 Very exaggerated/much too long; 2 a bit exaggerated/a bit too long; 3 just perfect; 4 a bit weak/a bit too short; 5 much too weak/much too short | Answers 1-3 = False; Answers 4-5 = True | Baseline |
| **HCW specifics** | | | | |
| Job: nurse | What is your job description? | Nurse; Medical doctors assistant; Physician; Secretary; Scientist; Physiotherapist; Ergotherapist; Logopedist; Social service; Dietician; Medical technical radiology assistant (MTRA); Housekeeping; Volunteer; Gastronomy/Hotel services; Kitchen; Technical service; Administration (Human resources, management); Informatics; Laboratory; other | Nurse = True; All others = False | Baseline |
| Job: physician | | | Physician = True; All others = False | Baseline |
| Full-time job (> 80%) | What is your work percentage? | Percentage (xxx%) | >80% = True; 0-80% = False | Baseline |
| Involved in AGP | In which aerosol-generating procedures have you been involved since March 2020? | None; In-/Extubation; Bronchoscopy; Reanimation; Tracheal secretion suction; Non-invasive ventilation; Gastroscopy; Transesophageal echocardiography; other | None = False; All others = True | Follow-up |
| Working in intensive case | In which department do you work? | List of departments | Intensive care unit = True, All others = False | Baseline |
| **Behaviour at work** | | | | |
| Hygiene knowledge | Which are standard hygiene measures? | Cough etiquette; Physical distancing*; Handhygiene; Vaccines; Wearing a gown when in contact with body fluids; Wearing surgical mask when having a cold (* incorrect answer, all others are correct) | Number of correct answers ≥ 3 = True; < 3 = False | Baseline |
| Regular meals in staff restaurant | How often do you go to the staff restaurant or the cafeteria? | None is available; Never; Less than weekly; Weekly, but not daily; Daily; More than daily; No restaurant or cafeteria visits | None, never or less than weekly; weekly or more | Baseline |
| Handwashing more frequent | Do you wash your hands more frequently in the current COVID-19 situation compared to before? | True; False | True; False | Baseline |
| **Use of personal protective equipment** | | | | |
| Always used goggles[1] | In what situations (during COVID-19 patient contact) did you use goggles? | 1 Never; 2 Occasionally; 3 Only if a contact with body fluids or secretions is expected; 4 Always | Answer 4 = True; Answers 1-3 = False | Follow-up |
| Always used gloves/gown[1] | In what situations (during COVID-19 patient contact) did you use gloves? In what situations (during COVID-19 patient contact) did you use a gown? | 1 Never; 2 Occasionally; 3 Only if a contact with body fluids or secretions is expected; 4 Always | Answer 4 (for both gloves and gown) = True; All others = False | Follow-up |
| Mostly used FFP2[1] | What mask do you usually wear in contact with COVID-19 patients, when non-aerosol generating procedures are done? Please consider the whole time since the start of the pandemic. | 1 Always surgical mask; 2 mostly surgical mask; 3 Both surgical mask and FFP2; 4 Mostly FFP2; 5 Always FFP2 | Answers 1-3 = False; Answers 4-5 = True | Follow-up |
| Always used FFP2 during AGP | What mask do you usually wear in contact with COVID-19 patients, when aerosol generating procedures are performed? Please consider the whole time since the start of the pandemic. | 1 Always surgical mask; 2 Mostly surgical mask; 3 Both surgical mask andFFP2; 4 Mostly FFP2; 5 Always FFP2 | Answers 1-4 = False; Answer 5 = True | Follow-up |

[1] In contact with COVID-19 patients outside of AGPs

Abbreviations: AU – Austria. GE – Germany. HCW – Health care worker. COPD – Chronic obstructive pulmonary disease. AGP – Aerosol-generating procedure. FFP2 – Filtering facepiece class 2.

Table S2. Cox regression analysis (full model and sensitivity analyses) with outcome “SARS-CoV-2- positive nasopharyngeal PCR/rapid antigen test”.

| | **Full model**[1] | | **Sensitivity analysis 1**[1] Excluding HCW with positive households | | **Sensitivity analysis 2** Treating cantons and institutions as fixed effects | | **Sensitivity analysis 3**[1] Restricting analysis to events after Dec 1st 2020 | |
|---|---|---|---|---|---|---|---|---|
| Participants; Events | n=3’259, n=433 | | n=2’783; n=250 | | n=3’259; n=433 | | n=3’067; n=203 | |
| **Risk or protection factor** | **HR (95% CI)** | **p** | **HR (95% CI)** | **p** | **HR (95% CI)** | **p** | **HR (95% CI)** | **p** |
| **Exposure to COVID-19** | | | | | | | | |
| Patient exposure *(time dependent)* | 2·24 (1·63-3·10) | <0·001 | 2·84 (2·05-3·93) | <0·001 | 2·11 (1·67-2·66) | <0·001 | 2·07 (1·55-2·76) | <0·001 |
| Coworker exposure *(time dependent)* | 1·58 (1·26-1·99) | <0·001 | 1·98 (1·58-2·48) | <0·001 | 1·56 (1·20-2·04) | 0·001 | 1·48 (1·13-1·94) | 0·004 |
| Household exposure *(time dependent)* | 10·06 (7·48-13·51) | <0·001 | NA | | 9·64 (7·66-12·12) | <0·001 | 14·21 (9·66-20·91) | <0·001 |
| Other exposure *(time dependent)* | 1·51 (1·31-1·73) | <0·001 | 1·70 (1·45-2·00) | <0·001 | 1·48 (1·20-1·83) | <0·001 | 1·64 (1·22-2·22) | 0·001 |
| Number of negative swabs *(time dependent)* | 0·91 (0·84-0·98) | 0·008 | 0·94 (0·83-1·05) | 0·277 | 0·91 (0·81-1·02) | 0·120 | 0·95 (0·87-1·04) | 0·243 |
| **Sociodemographic data** | | | | | | | | |
| Age (per 10 years) | 1·06 (1·00-1·13) | 0·046 | 0·97 (0·89-1·05) | 0·429 | 1·08 (0·98-1·18) | 0·133 | 0·96 (0·88-1·05) | 0·400 |
| Gender: male | 0·78 (0·61-1·01) | 0·058 | 0·50 (0·38-0·66) | <0·001 | 0·82 (0·60-1·11) | 0·191 | 0·64 (0·36-1·15) | 0·137 |
| Living in Germany or Austria | 1·10 (0·71-1·70) | 0·667 | 0·92 (0·40-2·11) | 0·837 | NA | | 0·69 (0·35-1·38) | 0·295 |
| Child in household | 1·13 (0·93-1·38) | 0·228 | 0·69 (0·55-0·87) | 0·002 | 1·10 (0·86-1·40) | 0·458 | 1·70 (1·25-2·29) | 0·001 |
| **Medical conditions** | | | | | | | | |
| Comorbidity | 0·99 (0·81-1·20) | 0·895 | 1·01 (0·76-1·33) | 0·965 | 1·00 (0·82-1·23) | 0·982 | 0·99 (0·72-1·34) | 0·923 |
| Active smoking | 0·65 (0·49-0·86) | 0·003 | 0·59 (0·41-0·85) | 0·005 | 0·66 (0·50-0·89) | 0·006 | 0·55 (0·29-1·06) | 0·074 |
| Pregnancy during study | 1·37 (1·05-1·77) | 0·019 | 1·27 (0·68-2·36) | 0·458 | 1·37 (0·87-2·16) | 0·172 | 2·03 (1·33-3·09) | 0·001 |
| **Behaviour outside of work** | | | | | | | | |
| Prophylactic home remedies | 1·08 (0·82-1·42) | 0·598 | 1·00 (0·76-1·30) | 0·974 | 1·07 (0·82-1·40) | 0·599 | 1·30 (0·94-1·78) | 0·107 |
| Social leisure activities | 0·93 (0·79-1·09) | 0·357 | 0·79 (0·67-0·94) | 0·007 | 0·91 (0·74-1·10) | 0·324 | 1·04 (0·79-1·37) | 0·781 |
| Wearing a mask outside work | 0·74 (0·56-0·99) | 0·040 | 0·69 (0·43-1·11) | 0·127 | 0·75 (0·58-0·97) | 0·026 | 0·84 (0·64-1·11) | 0·222 |
| Support for stronger public restrictions | 0·88 (0·73-1·05) | 0·161 | 0·92 (0·67-1·27) | 0·619 | 0·96 (0·73-1·25) | 0·749 | 1·11 (0·76-1·62) | 0·593 |
| **HCW specifics** | | | | | | | | |
| Job: nurse | 1·20 (1·03-1·39) | 0·020 | 1·43 (1·12-1·83) | 0·004 | 1·15 (0·89-1·49) | 0·276 | 1·24 (0·94-1·63) | 0·127 |
| Job: physician | 0·87 (0·65-1·15) | 0·328 | 1·10 (0·73-1·67) | 0·650 | 0·82 (0·58-1·15) | 0·256 | 0·90 (0·55-1·46) | 0·669 |
| Full-time job (> 80%) | 1·10 (0·94-1·27) | 0·230 | 1·10 (0·90-1·34) | 0·347 | 1·08 (0·85-1·39) | 0·521 | 1·51 (1·05-2·16) | 0·026 |
| Involved in AGP | 1·18 (0·96-1·45) | 0·114 | 1·21 (0·90-1·62) | 0·217 | 1·13 (0·91-1·40) | 0·266 | 1·31 (1·04-1·67) | 0·024 |
| Working in intensive care | 0·77 (0·53-1·12) | 0·170 | 0·63 (0·42-0·94) | 0·025 | 0·74 (0·50-1·10) | 0·142 | 1·02 (0·63-1·66) | 0·924 |
| **Behaviour at work** | | | | | | | | |
| Hygiene knowledge | 1·09 (0·91-1·32) | 0·342 | 1·01 (0·71-1·44) | 0·964 | 1·00 (0·75-1·34) | 0·976 | 1·30 (0·96-1·75) | 0·089 |
| Regular meals in staff restaurant | 1·16 (1·00-1·36) | 0·058 | 1·13 (0·95-1·34) | 0·173 | 1·15 (0·93-1·43) | 0·199 | 1·16 (0·84-1·59) | 0·373 |

| | | | | | | | | |
|---|---|---|---|---|---|---|---|---|
| **Use of personal protective equipment** | | | | | | | | |
| Always used goggles[2] | 0·81 (0·58-1·12) | 0·201 | 0·81 (0·53-1·25) | 0·342 | 0·88 (0·67-1·16) | 0·375 | 0·66 (0·40-1·08) | 0·100 |
| Always used gloves/gown[2] | 1·11 (0·71-1·73) | 0·644 | 1·11 (0·69-1·80) | 0·664 | 1·07 (0·81-1·41) | 0·636 | 1·16 (0·69-1·96) | 0·572 |
| **Mostly used FFP2**[2] | 0·80 (0·64-1·00) | 0·052 | 0·82 (0·54-1·25) | 0·360 | 0·88 (0·66-1·18) | 0·402 | 0·73 (0·55-0·97) | 0·029 |

[1] Institutions and living place (canton) included in model as random cluster term

[2] Germany/Austria treated like separate cantons and included as fixed effects

[3] In contact with COVID-19 patients outside of AGPs

Abbreviations: PCR – Polymerase chain reaction. HCW – Health care worker. AGP – Aerosol-generating procedure. FFP2 – Filtering facepiece class 2. NA – Not applicable

Table S3. Results of multivariable logistic regression analysis (full model and sensitivity analyses) regarding outcome "SARS-CoV-2 seroconversion".

| | **Full model**[1] | | **Sensitivity analysis 1**[1] Excluding HCW with positive households | | **Sensitivity analysis 2** Treating cantons and institutions as fixed effects | |
|---|---|---|---|---|---|---|
| Participants; Events | n=2916; n=511 | | n=2491; n=335 | | n=2916: n=511 | |
| **Risk or protection factor** | **OR (95% CI)** | **p** | **OR (95% CI)** | **p** | **OR (95% CI)** | **p** |
| **Sociodemographic data** | | | | | | |
| Age (per 10 years) | 0·96 (0·87 - 1·07) | 0·475 | 0·92 (0·82 - 1·04) | 0·177 | 0·96 (0·86 - 1·06) | 0·422 |
| Sex: male | 0·86 (0·62 - 1·18) | 0·349 | 0·77 (0·53 - 1·13) | 0·176 | 0·86 (0·62 - 1·19) | 0·368 |
| Living in Germany or Austria | 0·86 (0·47 - 1·58) | 0·628 | 0·81 (0·42 - 1·55) | 0·522 | NA | |
| No of additional persons in households | 0·88 (0·79 - 0·98) | 0·016 | 0·86 (0·76 - 0·98) | 0·022 | 0·87 (0·78 - 0·97) | 0·013 |
| Child in household | 0·97 (0·71 - 1·34) | 0·859 | 0·73 (0·48 - 1·10) | 0·132 | 0·98 (0·71 - 1·35) | 0·880 |
| Any positively tested household member | 5·01 (3·89 - 6·46) | <0·001 | NA | | 5·11 (3·96 - 6·60) | <0·001 |
| **Medical conditions** | | | | | | |
| Comorbidity | 0·93 (0·74 - 1·16) | 0·506 | 0·96 (0·74 - 1·24) | 0·732 | 0·93 (0·74 - 1·16) | 0·507 |
| Active smoking | 0·57 (0·41 - 0·79) | <0·001 | 0·56 (0·39 - 0·82) | 0·002 | 0·57 (0·41 - 0·78) | 0·001 |
| Pregnancy during study | 1·01 (0·56 - 1·82) | 0·979 | 0·91 (0·45 - 1·86) | 0·802 | 0·99 (0·53 - 1·74) | 0·969 |
| **Behaviour outside of work** | | | | | | |
| Prophylactic home remedies | 1·21 (0·90 - 1·63) | 0·205 | 1·14 (0·81 - 1·62) | 0·448 | 1·21 (0·89 - 1·63) | 0·211 |
| Social leisure activities | 1·02 (0·82 - 1·27) | 0·847 | 0·92 (0·72 - 1·18) | 0·517 | 1·02 (0·82 - 1·27) | 0·863 |
| Wearing a mask outside work | 0·96 (0·74 - 1·25) | 0·779 | 0·92 (0·68 - 1·24) | 0·588 | 0·97 (0·74 - 1·27) | 0·833 |
| Support for stronger public restrictions | 0·85 (0·64 - 1·14) | 0·282 | 0·93 (0·67 - 1·29) | 0·657 | 0·88 (0·65 - 1·17) | 0·386 |
| **HCW specifics** | | | | | | |
| Job: nurse | 1·55 (1·18 - 2·04) | 0·001 | 1·83 (1·33 - 2·52) | <0·001 | 1·56 (1·19 - 2·06) | 0·002 |
| Job: physician | 0·84 (0·59 - 1·21) | 0·360 | 0·88 (0·57 - 1·36) | 0·562 | 0·84 (0·58 - 1·20) | 0·337 |
| Full-time job (> 80%) | 1·08 (0·83 - 1·42) | 0·566 | 1·11 (0·81 - 1·51) | 0·521 | 1·10 (0·84 - 1·44) | 0·509 |
| Involved in AGP | 0·96 (0·75 - 1·24) | 0·775 | 1·03 (0·78 - 1·37) | 0·820 | 0·95 (0·74 - 1·21) | 0·659 |
| Working in ICU | 0·43 (0·27 - 0·68) | <0·001 | 0·40 (0·22 - 0·71) | 0·002 | 0·43 (0·27 - 0·68) | <0·001 |
| Frequency of contacts with COVID-19 patients | 1·51 (1·38 - 1·66) | <0·001 | 1·50 (1·35 - 1·68) | <0·001 | 1·51 (1·38 - 1·67) | <0·001 |
| Frequency of contacts with coworkers with COVID-19 | 1·22 (1·10 - 1·36) | <0·001 | 1·18 (1·05 - 1·33) | 0·007 | 1·22 (1·10 - 1·36) | <0·001 |
| **Behaviour at work** | | | | | | |
| Hygiene knowledge | 1·06 (0·77 - 1·46) | 0·698 | 1·12 (0·76 - 1·65) | 0·562 | 1·03 (0·75 - 1·44) | 0·834 |
| Regular meals in staff restaurant | 1·12 (0·88 - 1·41) | 0·363 | 1·06 (0·81 - 1·40) | 0·657 | 1·12 (0·88 - 1·42) | 0·362 |
| **Use of personal protective equipment** | | | | | | |
| Always used goggles[2] | 0·91 (0·68 - 1·22) | 0·521 | 0·99 (0·71 - 1·39) | 0·953 | 0·94 (0·70 - 1·26) | 0·677 |

| | | | | | | |
|---|---|---|---|---|---|---|
| Always used gloves/gown[2] | 0·81 (0·59 - 1·11) | 0·182 | 0·79 (0·55 - 1·13) | 0·197 | 0·80 (0·58 - 1·10) | 0·170 |
| **Mostly used FFP2**[2] | **0·73 (0·53 - 1·00)** | **0·053** | **0·69 (0·48 - 0·99)** | **0·046** | **0·76 (0·55 - 1·04)** | **0·088** |

[1]Institutions and living place (canton) included in model as random cluster term. For Sensitivity Analysis 2, the effects of these factors are not shown in the table.
[2] In contact with COVID-19 patients outside of AGPs
Abbreviations: HCW – Health care worker. AGP – Aerosol-generating procedure. FFP2 – Filtering facepiece class 2. NA – Not applicable.

Table S4. Subgroup analysis of HCW with frequent COVID-19 exposure *vs*. HCW with less frequent COVID-19 exposure; A) Cox regression (outcome SARS-CoV-2 positive swab); B) multivariable logistic regression (outcome SARS-CoV-2 seroconversion).

**A) Cox Regression**

| | **1−20 COVID-19 patient contacts** | | **>20 COVID-19 patient contacts** | |
|---|---|---|---|---|
| Participants; Events | n = 1292; n = 149 | | n = 1120; n = 215 | |
| **Risk or protection factor** | **HR** | **p** | **HR** | **p** |
| **Exposure to COVID-19** | | | | |
| Patient exposure *(time dependent)* | 2·09 (1·66-2·62) | <0·001 | 1·52 (0·93-2·49) | 0·099 |
| Coworker exposure *(time dependent)* | 1·64 (1·09-2·46) | 0·018 | 1·61 (0·99-2·63) | 0·055 |
| Household exposure *(time dependent)* | 13·93 (9·64-20·12) | <0·001 | 6·49 (4·02-10·47) | <0·001 |
| Other exposure *(time dependent)* | 1·71 (1·36-2·14) | <0·001 | 1·20 (0·86-1·66) | 0·286 |
| Number of negative swabs *(time dependent)* | 0·88 (0·76-1·01) | 0·060 | 0·93 (0·83-1·03) | 0·143 |
| **Sociodemographic data** | | | | |
| Age (per 10 years) | 1·10 (0·99-1·22) | 0·086 | 1·05 (0·95-1·15) | 0·338 |
| Sex:male | 0·53 (0·29-0·98) | 0·042 | 0·86 (0·63-1·17) | 0·334 |
| Living in Germany or Austria | 1·20 (0·35-4·06) | 0·775 | 1·11 (0·49-2·52) | 0·794 |
| Child in household | 0·93 (0·65-1·34) | 0·710 | 1·24 (0·84-1·84) | 0·283 |
| **Medical conditions** | | | | |
| Comorbidity | 0·95 (0·71-1·27) | 0·705 | 0·99 (0·72-1·35) | 0·938 |
| Active smoking | 0·72 (0·38-1·36) | 0·307 | 0·61 (0·32-1·14) | 0·120 |
| Pregnancy during study | 1·41 (0·87-2·29) | 0·165 | 0·88 (0·40-1·94) | 0·745 |
| **Behaviour outside of work** | | | | |
| Prophylactic home remedies | 1·02 (0·70-1·49) | 0·909 | 1·09 (0·76-1·58) | 0·640 |
| Social leisure activities | 1·03 (0·76-1·41) | 0·836 | 0·91 (0·73-1·12) | 0·373 |
| Wearing a mask outside work | 0·70 (0·47-1·04) | 0·078 | 0·82 (0·61-1·11) | 0·193 |
| Support for stronger public restrictions | 0·91 (0·57-1·48) | 0·715 | 0·81 (0·65-1·02) | 0·076 |
| **HCW specifics** | | | | |
| Job: nurse | 1·20 (0·74-1·94) | 0·450 | 1·77 (1·30-2·41) | <0·001 |
| Job: physician | 0·53 (0·30-0·94) | 0·029 | 1·48 (0·91-2·39) | 0·111 |
| Full-time job (> 80%) | 1·10 (0·79-1·53) | 0·561 | 1·09 (0·85-1·39) | 0·504 |
| Involved in AGP | 1·08 (0·88-1·34) | 0·453 | 1·12 (0·76-1·65) | 0·566 |
| Working in intensive care | 0·64 (0·35-1·17) | 0·149 | 0·67 (0·45-0·99) | 0·046 |
| **Behaviour at work** | | | | |
| Hygiene knowledge | 1·12 (0·80-1·57) | 0·521 | 1·07 (0·78-1·46) | 0·691 |
| Regular meals in staff restaurant | 1·28 (0·89-1·84) | 0·186 | 1·02 (0·80-1·30) | 0·879 |
| **Use of personal protective equipment** | | | | |
| Always goggles[1] | 0·76 (0·45-1·29) | 0·309 | 0·93 (0·68-1·27) | 0·664 |

| | | | | |
|---|---|---|---|---|
| Always gloves/gown[1] | 0·82 (0·42-1·61) | 0·562 | 1·28 (0·92-1·78) | 0·147 |
| **Mostly using FFP2 mask**[1] | 1·06 (0·70-1·63) | 0·774 | 0·66 (0·54-0·81) | <0·001 |

**B) Multivariable Logistic Regression**

| | **1−20 COVID-19 patient contacts** | | **> 20 COVID-19 patient contacts** | |
|---|---|---|---|---|
| Participants; Events | n = 1156; n = 164 | | n = 1019; n = 272 | |
| **Risk or protection factor** | OR (95% CI) | p | OR (95% CI) | p |
| **Exposure to COVID-19** | | | | |
| Frequency of contacts to coworkers with COVID-19 | 1·22 (1·02 - 1·45) | 0·032 | 1·23 (1·04 - 1·45) | 0·016 |
| Any positively tested household member | 3·97 (2·60 - 6·06) | <0·001 | 5·28 (3·59 - 7·78) | <0·001 |
| **Sociodemographic data** | | | | |
| Age (per 10 years) | 1·03 (0·86 - 1·22) | 0·776 | 0·91 (0·78 - 1·06) | 0·233 |
| Sex:male | 0·66 (0·37 - 1·20) | 0·175 | 0·95 (0·62 - 1·47) | 0·822 |
| Living in Germany or Austria | 1·28 (0·51 - 3·22) | 0·606 | 0·74 (0·35 - 1·57) | 0·427 |
| Number of additional persons in household | 0·94 (0·78 - 1·14) | 0·536 | 0·86 (0·74 - 1·01) | 0·066 |
| Child in household | 0·81 (0·46 - 1·41) | 0·450 | 1·08 (0·67 - 1·72) | 0·759 |
| **Medical conditions** | | | | |
| Comorbidity | 0·76 (0·52 - 1·13) | 0·173 | 1·02 (0·74 - 1·42) | 0·890 |
| Active smoking | 0·76 (0·46 - 1·27) | 0·296 | 0·49 (0·30 - 0·80) | 0·004 |
| Pregnancy during study | 1·13 (0·47 - 2·75) | 0·781 | 0·79 (0·28 - 2·24) | 0·657 |
| **Behaviour outside of work** | | | | |
| Prophylactic home remedies | 1·29 (0·80 - 2·10) | 0·298 | 1·11 (0·69 - 1·80) | 0·665 |
| Social leisure activities | 1·02 (0·71 - 1·46) | 0·931 | 1·10 (0·80 - 1·52) | 0·545 |
| Wearing a mask outside work | 0·82 (0·51 - 1·29) | 0·388 | 1·18 (0·80 - 1·74) | 0·404 |
| Support for stronger public restrictions | 1·01 (0·62 - 1·66) | 0·970 | 0·83 (0·54 - 1·27) | 0·395 |
| **HCW specifics** | | | | |
| Job: nurse | 1·39 (0·89 - 2·17) | 0·151 | 2·22 (1·40 - 3·52) | <0·001 |
| Job: physician | 0·50 (0·26 - 0·96) | 0·038 | 1·33 (0·78 - 2·28) | 0·298 |
| Full-time job (> 80%) | 1·31 (0·82 - 2·08) | 0·254 | 0·97 (0·64 - 1·48) | 0·901 |
| Involved in AGP | 1·35 (0·90 - 2·03) | 0·144 | 0·82 (0·58 - 1·15) | 0·251 |
| Working in intensive care | 0·33 (0·11 - 1·00) | 0·049 | 0·51 (0·30 - 0·87) | 0·015 |
| **Behaviour at work** | | | | |
| Hygiene knowledge | 0·99 (0·59 - 1·65) | 0·968 | 1·12 (0·65 - 1·93) | 0·691 |
| Regular meals in staff restaurant | 1·53 (1·03 - 2·28) | 0·037 | 1·03 (0·72 - 1·47) | 0·886 |
| **Use of personal protective equipment** | | | | |
| Always goggles[1] | 0·90 (0·54 - 1·51) | 0·703 | 1·06 (0·72 - 1·58) | 0·755 |
| Always gloves/gown[1] | 0·60 (0·35 - 1·03) | 0·065 | 0·98 (0·64 - 1·49) | 0·915 |
| **Mostly using FFP2 mask**[1] | **0·75 (0·42 - 1·32)** | **0·320** | **0·64 (0·42 - 0·97)** | **0·036** |

[1] In contact with COVID-19 patients outside of AGPs

Abbreviations: HCW – Health care worker. AGP – Aerosol-generating procedure. FFP2 – Filtering facepiece class 2. NA – not applicable

Table S5. Subgroup analysis of HCW performing AGP: A) Cox regression (outcome SARS-CoV-2 positive swab); B) multivariable logistic regression (outcome SARS-CoV-2 seroconversion).

| A) Cox Regression | Participants involved in AGP | |
|---|---|---|
| Participants; Events | n = 1204; n = 188 | |
| **Risk or protection factor** | **HR** | **p** |
| **Exposure to COVID-19** | | |
| Patient exposure *(time dependent)* | 1·50 (0·98-2·32) | 0·065 |
| Coworker exposure *(time dependent)* | 1·72 (1·24-2·37) | 0·001 |
| Household exposure *(time dependent)* | 9·75 (6·28-15·14) | <0·001 |
| Other exposure *(time dependent)* | 1·42 (1·07-1·88) | 0·015 |
| Number of negative swabs *(time dependent)* | 0·88 (0·75-1·03) | 0·108 |
| **Sociodemographic data** | | |
| Age (per 10 years) | 1·02 (0·88-1·19) | 0·776 |
| Sex:male | 0·87 (0·57-1·33) | 0·531 |
| Living in Germany or Austria | 1·46 (0·79-2·67) | 0·224 |
| Child in household | 1·15 (0·88-1·50) | 0·297 |
| **Medical conditions** | | |
| Comorbidity | 1·13 (0·85-1·49) | 0·402 |
| Active smoking | 0·65 (0·37-1·13) | 0·129 |
| Pregnancy during study | 0·56 (0·13-2·38) | 0·433 |
| **Behaviour outside of work** | | |
| Prophylactic home remedies | 1·11 (0·67-1·83) | 0·685 |
| Social leisure activities | 0·98 (0·77-1·26) | 0·900 |
| Wearing a mask outside work | 0·75 (0·51-1·11) | 0·157 |
| Support for stronger public restrictions | 0·91 (0·69-1·20) | 0·512 |
| **HCW specifics** | | |
| Job: nurse | 1·17 (0·87-1·58) | 0·309 |
| Job: physician | 1·12 (0·64-1·96) | 0·697 |
| Full-time job (> 80%) | 0·95 (0·65-1·37) | 0·767 |
| Working in intensive care | 0·77 (0·48-1·24) | 0·277 |
| **Behaviour at work** | | |
| Hygiene knowledge | 1·72 (1·03-2·88) | 0·039 |
| Regular meals in staff restaurant | 1·00 (0·67-1·47) | 0·981 |
| **Use of personal protective equipment** | | |
| Always goggles[1] | 0·78 (0·59-1·03) | 0·075 |
| Always gloves/gown[1] | 1·11 (0·67-1·82) | 0·686 |
| **Mostly using FFP2 mask[1]** | **0·71 (0·50-1·01)** | **0·059** |

| | | |
|---|---|---|
| Always using FFP2 mask during AGP | 1·08 (0·71-1·64) | 0·726 |

**B) Multivariable Logistic Regression**

| | Participants involved in AGP | |
|---|---|---|
| Participants; Events | n = 1087; n = 216 | |
| **Risk or protection factor** | **OR (95% CI)** | **p** |
| **Exposure to COVID-19** | | |
| Frequency of contacts to COVID-19 patients | 1·44 (1·22 - 1·70) | <0·001 |
| Frequency of contacts to coworkers with COVID-19 | 1·27 (1·06 - 1·51) | 0·008 |
| Any positively tested household member | 4·69 (3·14 - 7·00) | <0·001 |
| **Sociodemographic data** | | |
| Age (per 10 years) | 1·01 (0·85 - 1·19) | 0·951 |
| Sex:male | 0·99 (0·64 - 1·53) | 0·953 |
| Living in Germany or Austria | 1·38 (0·65 - 2·91) | 0·404 |
| Number of additional persons in households | 0·89 (0·75 - 1·06) | 0·185 |
| Child in household | 0·90 (0·53 - 1·52) | 0·693 |
| **Medical conditions** | | |
| Comorbidity | 1·08 (0·76 - 1·52) | 0·667 |
| Active smoking | 0·46 (0·28 - 0·78) | 0·004 |
| Pregnancy during study | 1·25 (0·39 - 3·95) | 0·708 |
| **Behaviour outside of work** | | |
| Prophylactic home remedies | 1·41 (0·88 - 2·25) | 0·149 |
| Social leisure activities | 1·05 (0·75 - 1·47) | 0·786 |
| Wearing a mask outside work | 0·84 (0·56 - 1·26) | 0·404 |
| Support for stronger public restrictions | 1·11 (0·73 - 1·69) | 0·633 |
| **HCW specifics** | | |
| Job: nurse | 1·28 (0·78 - 2·09) | 0·330 |
| Job: physician | 0·97 (0·55 - 1·73) | 0·930 |
| Full-time job (> 80%) | 0·95 (0·62 - 1·46) | 0·822 |
| Working in intensive care | 0·44 (0·27 - 0·74) | 0·002 |
| **Behaviour at work** | | |
| Hygiene knowledge | 1·81 (0·94 - 3·50) | 0·077 |
| Regular meals in staff restaurant | 1·09 (0·75 - 1·59) | 0·645 |
| **Use of personal protective equipment** | | |
| Always used goggles[1] | 0·89 (0·59 - 1·32) | 0·552 |
| Always used gloves/gown[1] | 0·86 (0·56 - 1·34) | 0·518 |
| **Mostly used FFP2 mask[1]** | **0·66 (0·43 - 1·01)** | **0·055** |
| Always used FFP2 mask during AGP | 0·89 (0·62 - 1·28) | 0·535 |

[1] In contact with COVID-19 patients outside of AGPs

Abbreviations: HCW – Health care worker. AGP – Aerosol-generating procedure. FFP2 – Filtering facepiece class 2. NA – not applicable
